# What does it take to make progress in a disease?

**DOI:** 10.1101/2024.02.27.24303441

**Authors:** Michael S. Ringel, Julie Dethier, Michelle J. Davitt, Maria Denslow, R. Andrew Fowler, Sebastian C. Hasenfuss, Ulrik Schulze

## Abstract

In this paper, we investigate what conditions need to be in place to make progress in combating a disease using a case-control design: we compare cases (diseases with a successful therapy) to controls (diseases without a successful therapy). We find five conditions (“hurdles”) must typically be cleared for success: (A) understanding of biological drivers, (B) ability to modulate biology, (C) availability of translational models, (D1) ability to identify patients, and (D2) ability to measure clinical response. This framework is similar to ones deployed to evaluate individual drug candidates but is employed here to make inferences about entire diseases. It can be used to identify diseases most ready for progress, where efforts should be focused to make progress in diseases that are currently intractable, and where the industry could benefit from development of tools to address the hurdle that is most commonly the last to be cleared across diseases—namely, (C) translational models.

## Introduction

Biopharmaceutical research and development (R&D) is a difficult, lengthy, expensive, and risky process.^1,2,3,4,5,6^ In particular, despite a recent resurgence in R&D success rates,^7,8,9^ the vast majority of R&D projects ultimately fail, resulting in lost opportunity to bring new medicines to patients and wasted expense and effort.^10,11,12,13,14^ The most effective single lever for improving R&D productivity is to reduce this cost of failure, either by increasing overall success rates or by shifting failures earlier, thus freeing up capacity to be deployed to other, higher-probability projects.^6,15,16^

A number of biopharmaceutical companies have explored ways to improve their decision-making regarding which R&D projects to continue and which to terminate, by increasing the quality of information available to decision-makers, the quality of the decision-making process itself, or both.^17,18,19^ In particular, a number of companies have laudably introduced asset-level frameworks that require validation that scientists are pursuing the “right biological target” to have an effect on the disease, using the “right molecule” to engage the target both effectively and safely, testing the hypothesis in the “right patients” to see an effect, and so on.^20,21,22^ It is plausible that these efforts, coupled with a significant increase in known disease targets with human validation via “omics tools,”^23,24,25^ are behind the recent improvements in R&D success rates and cost of failure.^7^

This framework regarding predictors of success probability has generally been applied at the level of individual drug candidates. In this paper, we argue that the framework also works well at a level above drug candidates: entire diseases. That is, one can predict whether success in fighting a disease is likely by applying this framework. We provide evidence for this assertion by conducting a “case-control” study of “cases” (diseases for which a successful therapy has been developed) versus “controls” (diseases without a successful therapy) and the association between these states and the status of five predictor variables (“hurdles” to overcome, described in more detail ahead). Moreover, not only does this framework provide a lens into whether progress in combating a disease is likely, but it also provides a guide as to which hurdles should be the focus of attention in order to address a currently intractable disease. Lastly, by examining which hurdles show up most frequently as barriers to progress across multiple diseases, this framework provides a guide for industry and academia on what efforts might be helpful to improve the chances of success more broadly across all diseases—the missing link in drug R&D.

### Framework

The events necessary to discover and develop a drug are typically divided into a step-wise sequence starting with discovery biology followed by lead generation and optimization, preclinical testing, and ultimately clinical validation and submission to regulators (see exhibit 1). There is no absolute reason the activities of drug R&D need to be linear or follow exactly this order, and indeed there are many instances in which it has not followed the archetypal sequence. For example, many drug development efforts in oncology and rare disease have employed a hybrid approach that goes straight from Phase I safety testing in patients to Phase III trials.^26,27^ Furthermore, the history of drug R&D is replete with examples of phenotypic discovery for which targets were never identified,^28,29,30,31,32^ or for which the sequence of activities happened in virtually every permutation of the archetype.^33^ Lastly, the exact tools used today need not remain the tools used going forward: for instance, while preclinical evidence generation has historically relied on animal models and biochemical assays, these have proven to be poor predictors of human-organism-level responses.^34,35,36^ As a result, there is an emerging potential to augment or replace these tools with human genetic evidence^23,25^ and organoid, organ-on-a-chip, or in-silico methods.^36,37,38,39,40,41,42^

This being said, at a conceptual level, there are still four key questions that must be answered, in whatever sequence and manner, to develop a new therapy, with the caveats mentioned previously. These questions are: i) Is there an aspect of biology available that can affect disease pathophysiology? ii) How can that biology be modulated? iii) What evidence is sufficient to allow testing this hypothesis in humans? and iv) What evidence is sufficient to enable a regulator to approve clinical use in patients? Based on this, we hypothesized that there are five hurdles that generally need to be cleared for success in a disease area (as illustrated in exhibit 1): (A) understanding of biological drivers, (B) ability to modulate biology, (C) availability of translational models, (D1) ability to identify patients, and (D2) ability to measure clinical response—with the final two hurdles being related aspects of successful clinical development.

**Exhibit 1:**
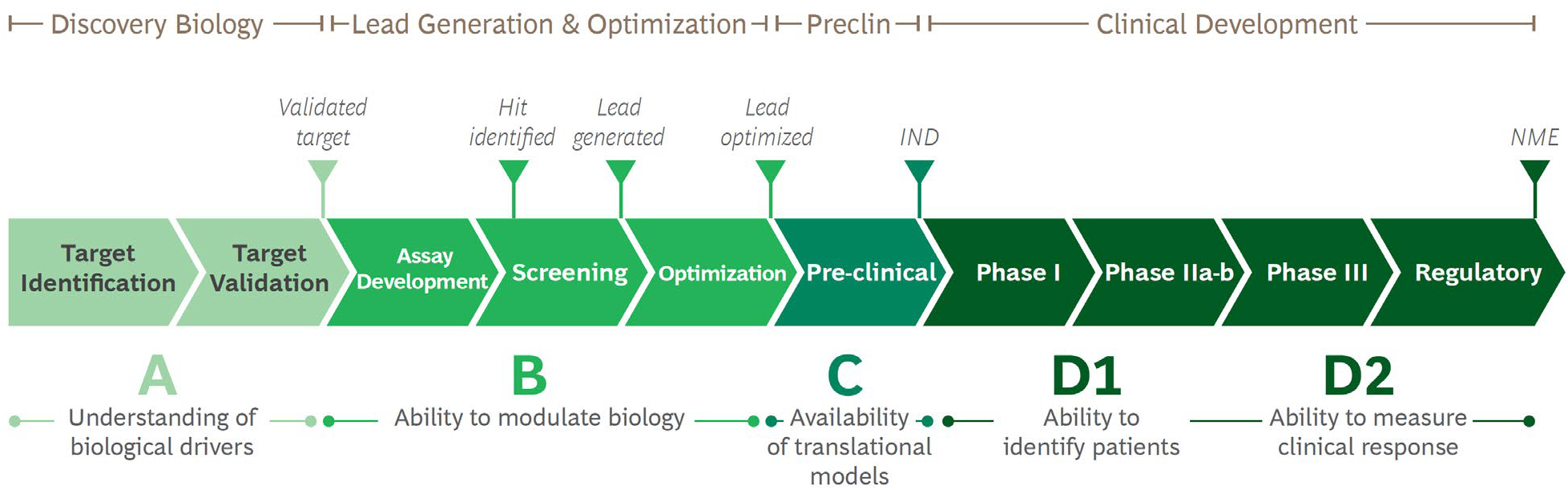
Steps in drug R&D with the five hurdles highlighted. This archetypal sequence of activities covers four main steps: discovery biology to elucidate and validate biological drivers of interest, lead generation and optimization to determine how to modulate the biology of interest, preclinical testing of safety and efficacy, and clinical testing in humans, typically in a stepwise fashion covering Phase I, safety testing in normal, healthy volunteers, Phase IIa dose determination, Phase IIb clinical proof-of-concept, and Phase III registrational studies. Each of these four steps corresponds to the five hurdles to clear for success in a disease area, with clinical testing in humans having two hurdles: ability to identify patients and measure clinical response. IND = investigational new drug filing, NME = new molecular entity approval by regulatory authorities.

## Methods

We compared “cases” (diseases that have a successful therapy) against “controls” (diseases that do not have a successful therapy) across disease areas to identify patterns common to diseases with successful therapies versus without. We used 16 disease areas as defined by Anatomical Therapeutic Chemical (ATC) classification codes, and selected two high-prevalence cases and two high-prevalence controls within each disease area for a total of 64 diseases analyzed. We excluded diseases that were not defined clearly enough, clinical presentations of many disease types, descriptions of procedures, and conditions caused by malnutrition or poisoning (see exhibit 2). We defined success as the existence of an approved drug therapy with significant impact on clinical outcomes. We restricted our analysis to the time period prior to the COVID-19 pandemic to avoid potential transient effects (e.g., disruption of clinical trials) that are not expected to persist post-pandemic.

**Exhibit 2:**
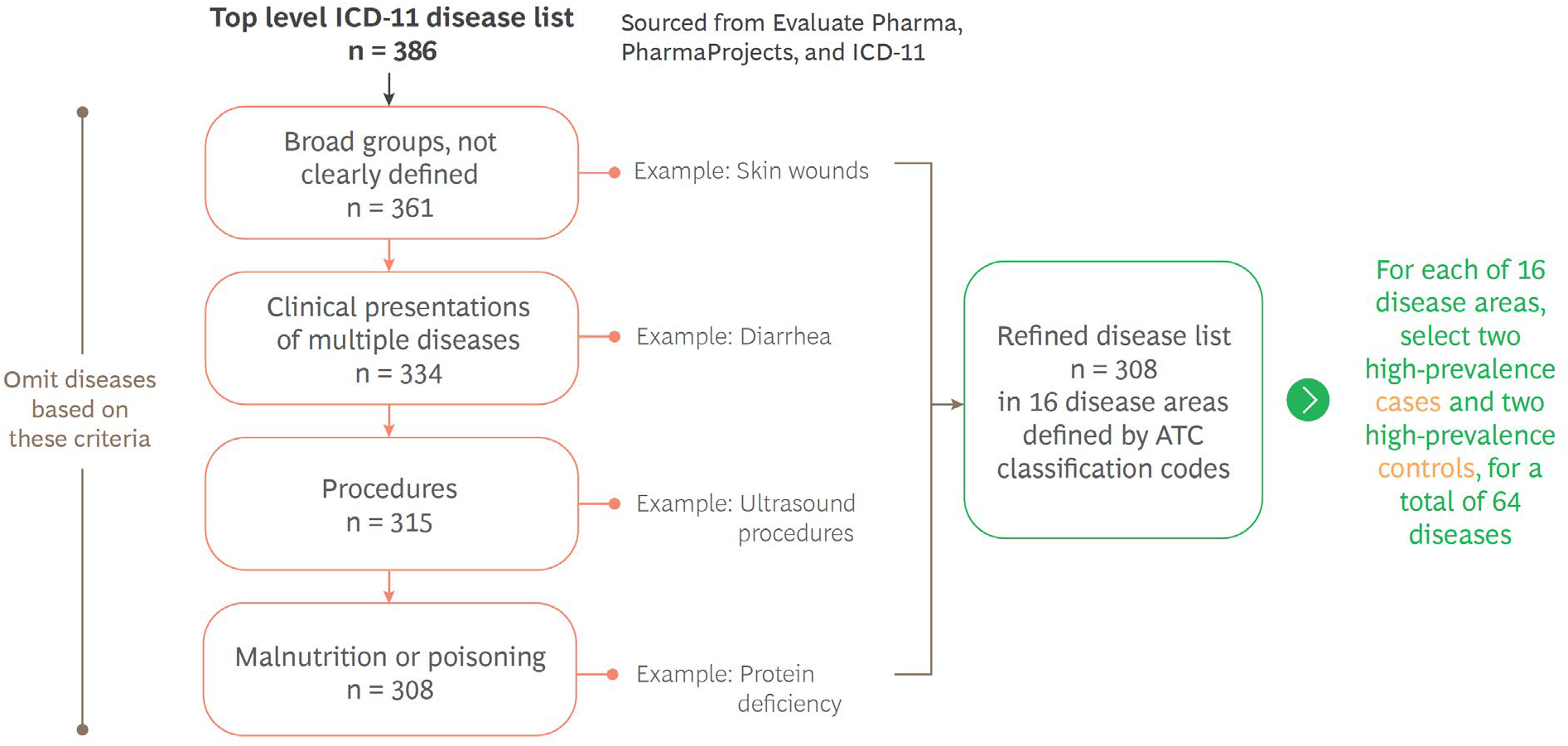
Selection scheme for included diseases. The cases and controls for the analysis were obtained by selecting two cases and two controls each with high prevalence in the US from each of 16 disease areas, filtering diseases that fit one or more of the criteria in the exhibit. ICD-11: international classification of disease, revision 11. ATC: anatomical therapeutic chemical.

For the selected diseases, we then scored achievement against each of the five hurdles, scoring two ways: First, we asked a set of 18 physicians each independently and blindly (i.e., agnostic of the hypothesis being tested) to score each disease. Second, we performed our own literature analysis to score the diseases, in particular paying attention to the clearance of the hurdles prior to the launch of therapy, and found similar results to the physician-driven approach.

Each of the five hurdles was scored using a rubric that defined a four-point scale (0 to 3), with one point awarded for each of three elements that could be true for each hurdle, as described below.

For hurdle (A), understanding of biological drivers, the elements were the following:

- Disease driver has been identified (genetically or otherwise).
- Strong gene-disease association exists (clinical and supporting experimental evidence provided by multiple studies).
- Many single-nucleotide polymorphisms are associated with the gene-disease pair, or there is a well-known mechanism of action for non-genetic drivers.

For hurdle (B), ability to modulate biology, the elements were the following:

- Target class is accessible (e.g., experience in target class, or known manner of access).
- Target class has been successfully targeted in the past.
- Organ/cell type easily accessible (e.g., does not need to cross the blood-brain barrier).

For hurdle (C), availability of translational models, the elements were the following:

- Models have same driver (gene/mutation or other) as in human disease.
- Models have similar in vivo context to human disease (e.g., chronic vs. acute, immunosuppressed, specific diet).
- Models display a phenotype similar to that of human disease.

For hurdle (D1), ability to identify patients, the elements were the following:

- A diagnostic exists that is specific and sensitive.
- The diagnostic is minimally invasive, safe, inexpensive, and easy to perform (e.g., blood/ urine test, physical exam, questionnaire, ultrasound or x-rays versus biopsy, endoscopy, CT scan, or MRI).
- Robust biomarker(s) allows patient stratification.

For hurdle (D2), ability to measure clinical response, the elements were the following:

- A test exists that is specific and sensitive.
- The test is minimally-invasive, safe, inexpensive, and easy to perform (e.g., blood/urine test, physical exam, questionnaire, ultrasound or x-rays versus biopsy, endoscopy, CT scan, or MRI).
- The test allows real-time updates and a dynamic view of patient state.

Each hurdle was considered cleared for a disease if the score was greater than or equal to 2 points (i.e., at least two elements were true). For hurdle (B), ability to modulate biology, no score was assigned if the target was not known. Timing of hurdle clearance was determined based on a literature review and timing of a therapy on the approval or marketing start date for the drug, and in our literature review-based scoring, we only included clearance of a hurdle if it preceded the achievement of the therapy approval or marketing start date. To estimate the venture capital (VC) investment against each hurdle, we grouped companies with VC funding into nine categories: Target, Modality, Platform, Model, Diagnostic, Device, Drug delivery, Computational/Digital, and R&D model. We then computed the proportion of companies in each category.

## Results

### Clearing the hurdles independently and collectively correlates with progress in a disease

In comparing cases versus controls, we find that clearance of the hurdles does correlate with progress in developing therapies for diseases. In one-way ANOVA comparisons, each of the hurdles shows a statistically significant correlation with the development of the therapy, except for hurdle (A), understanding of biological drivers. Furthermore, the five-way interaction term for achievement of all hurdles is also significant, indicating that success is most likely achieved when all hurdles are cleared (see table 1). Using a random forest model, we find strong predictive power with an area under the receiver operating characteristic curve of 0.91; that is, for a false positive rate of 11%, 85% of true positives were identified in the test set.^43,44^

**Table 1:**
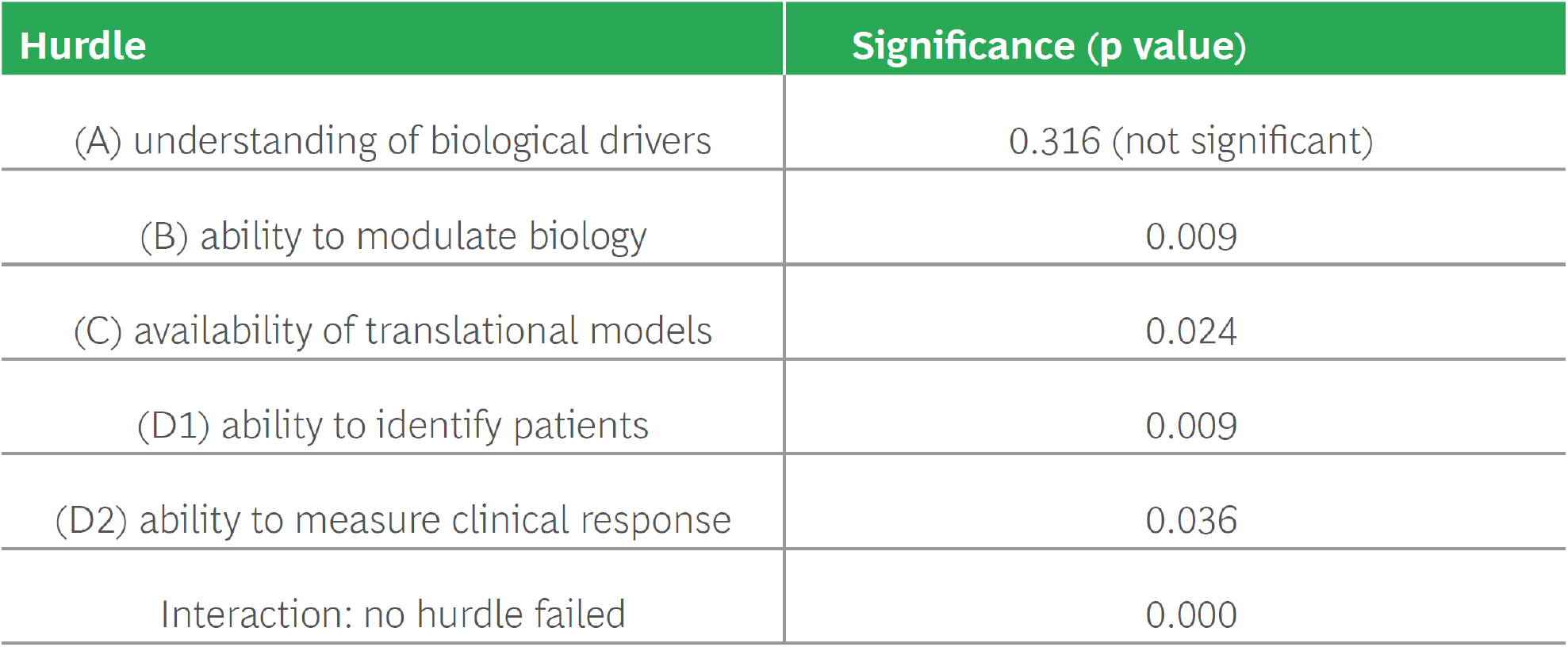
Results of one-way ANOVA. In one-way tests of the correlation of progress in combating a disease with each of the variables independently as well as an interaction term that is true only if no hurdle has failed to be cleared, each of the correlations is significant except hurdle (A)—see text for full explanation.

| Hurdle | Significance (p value) |
| --- | --- |
| (A) understanding of biological drivers | 0.316 (not significant) |
| (B) ability to modulate biology | 0.009 |
| (C) availability of translational models | 0.024 |
| (D1) ability to identify patients | 0.009 |
| (D2) ability to measure clinical response | 0.036 |
| Interaction: no hurdle failed | 0.000 |

Hurdle (A), understanding of biological drivers, was not significant in our analysis. This is not entirely surprising given the many times phenotypic drug development without target understanding has been successful.^28,29,30,31,32,33^ However, there is a trend toward greater understanding of biology in cases versus controls (see exhibit 3), which we interpret as clearing hurdle (A) being not an absolute condition precedent but still valuable in many situations; that is, there is merit in both target-based and phenotypic screening approaches. We see evidence for this in the fact that the interaction effect across all hurdles is significant. We suspect that, with a larger sample size, we would be able to discriminate a direct signal for hurdle (A).

Hurdle (B), ability to modulate biology, does correlate with success; however, it is increasingly rare to find situations in which the biology is inaccessible to modulation. Over the last several decades, there has been a profusion of new modalities able to access virtually all target space,^45,46^ as well as advances in approaches for traditional modalities such as structure-based drug design (SBDD) that have improved the ability of these modalities to modulate previously intractable targets.^47,48,49^ Although out of scope for this analysis, the response to the COVID-19 pandemic is an instructive example, with both new modalities such as mRNA^50,51^ and viral vectors,^52,53^ by-now-established modalities such as monoclonal antibodies,^54,55,56,57,58^ and more-established modalities such as protein subunits^59^ and small molecules^60,61,62,63^ (some enabled by SBDD^64^) all playing a role. This expansion of our therapeutic armamentarium has led to novel therapies for a variety of diseases, including many cancers and autoimmune disorders such as multiple sclerosis, asthma, and rheumatoid arthritis.^65,66,67,68,69^

**Exhibit 3:**
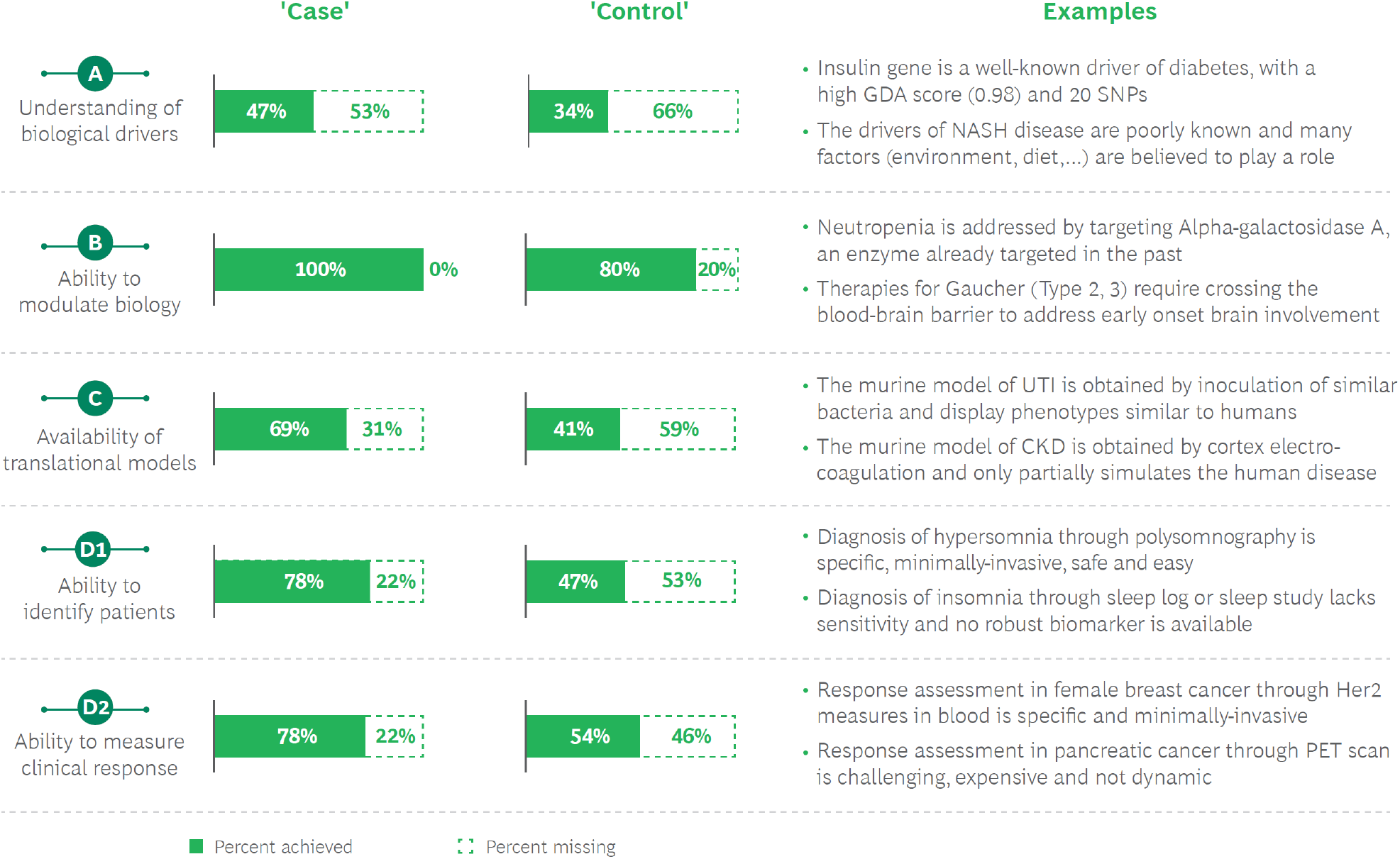
Correlations of hurdles with development of successful therapy. Each of the hurdles is more frequently cleared for cases versus controls (though not statistically significant for hurdle (A)). GDA: gene-disease association, NASH: non-alcoholic steatohepatitis, UTI: urinary tract infection, CKD: chronic kidney disease, PET: positron emission tomography.

Hurdle (C), availability of translational models, has a low level of achievement for both cases (69%) and controls (41%). Critically, however, it is most often the last hurdle to be cleared— true in 50% of all cases (see exhibit 4). The availability of a good translational model has often been the factor unleashing the ability to innovate (a topic to which we return in the Discussion section).

Lastly, hurdles (D1) ability to identify patients and (D2) ability to measure clinical response are the hurdles with the largest difference between cases and controls, true in nearly 80% of cases but only 47% and 54% of controls, respectively.

**Exhibit 4:**
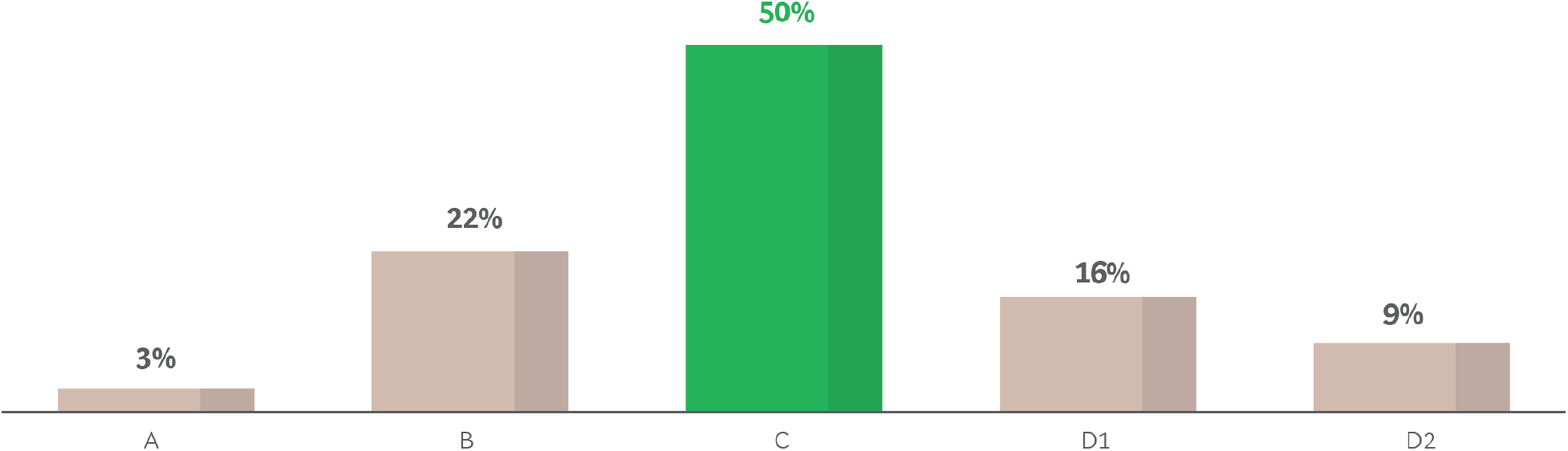
“Last hurdle to be cleared” among cases. For cases (diseases with successful treatment), the last hurdle to be cleared (when including only hurdles cleared pre-approval) is most frequently hurdle (C), availability of translational models.

## Discussion

### Inferences about diseases ready for progress

Our model allows for predictive analysis. At the time we conducted the analysis, there were several diseases in our control set with four of the five hurdles cleared. We would predict that these diseases are more ripe for progress. Indeed, this has since proven to be the case for one of the diseases in our control set: Duchenne Muscular Dystrophy (DMD). The genetic driver of the disease was well-known: mutations in the DMD gene alter the structure or function of dystrophin or prevent any functional dystrophin from being produced.^70,71,72^ Available translational models (e.g., the X chromosome-linked muscular dystrophy (mdx) mouse)^73^ provided a valuable model for DMD in humans, albeit with a milder muscle-wasting phenotype than in humans.^71^ Diagnosis for DMD was specific and sensitive thanks to genetic testing and muscle biopsy. In addition, plasma creatine kinase level, which is expressed in muscle and brain cells, offered a robust biomarker for the disease.^71,72^ Yet, at the time of the analysis, limited therapeutic options were available for treatment of DMD,^70,71^ with glucocorticoids the mainstay of treatment. Sarepta’s Exondys 51 (eteplirsen), based on exon-skipping technology, was conditionally approved by the U.S. Food and Drug Administration (FDA) in 2016 for DMD patients with mutations in the dystrophin gene amenable to exon 51 skipping.^74^ Yet, in 2018, the Committee for Medicinal Products for Human Use of the European Medicines Agency issued a negative opinion of eteplirsen for the treatment of DMD.^75^ But since 2019, three more therapies for DMD have been approved by the FDA: Sarepta’s Vyondys 53 (golodirsen) for exon 53 skipping in December 2019,^76^ Nippon Shinyaku’s Viltepso (viltolarsen) for exon 53 skipping in August 2020 (also approved in Japan),^77^ and Sarepta’s Amondys 45 (casimersen) for exon 45 skipping in February 2021.^78^

Taking a snapshot at the present time, we can see other diseases that have currently cleared four or five of the hurdles but do not currently have an approved therapy with a significant clinical benefit. An example is hereditary angioedema (HAE). The genetic drivers of the disease are well-known: mutations of the C1NH/SERPING1 gene encoding the C1 inhibitor (C1INH) lead to plasma deficiency and recurrent attacks of severe swelling.^79^ There are murine models that share the same disease etiology (disruption of the C1INH gene)^80^ and the same clinical features (acute hereditary angioedema attacks).^81^ Patients can be identified by specific and sensitive laboratory tests (biomarker measurement of serum antigenic C4) confirmed by screening for the SERPING1 or C1NH genes.^82,83^ Yet, therapeutic options are currently limited, focused on symptom management and not long-term prophylaxis (LTP).^84,85^ New drugs are being investigated for LTP. Some, such as KalVista’s sebetralstat, exert an anti-plasma K action to inhibit the kallikrein-kinin system.^86^ Others, such as Ionis’s donidalorsen, act by inhibiting prekallikrein, with a subsequent decrease in plasma K.^87^ Still others, such as CSL’s garadacimab, block factor XII.^88^ Our framework suggests that at least one of these therapies should prove successful.

### Actions to make progress in a currently intractable disease

Contrary to the situation with DMD and HAE, there are many diseases for which a number of the conditions for progress are currently missing. This is not a call to eschew R&D in these areas; instead, it can be a roadmap for the actions most necessary to make progress by focusing attention on the key gaps. Per the analysis in exhibit 4, often the key gaps to be resolved are (C) availability of translational models and (D1) ability to identify patients with suitable biomarkers, as well as (perhaps more historically, given many recent advances in modalities) (B) ability to modulate biology. One pertinent example is nonalcoholic steatohepatitis (NASH). Experts are not sure why some people with non-alcoholic fatty liver disease have NASH while others have simple fatty liver.^89,90^ Available models, although providing critical guidance in understanding specific stages of the disease pathogenesis and progression, require further development to better mimic the disease spectrum in order to provide both increased mechanistic understanding and identification/testing of novel therapeutic approaches.^91,92,93^ Specific diagnosis is challenging and liver biopsy remains the gold standard for definitive diagnosis.^94,95^

### Patterns across diseases

The last kind of inference that can be made from our findings is one regarding patterns across diseases. Are there certain keys to success that can be gleaned from our analysis that might point to untapped areas of opportunity?

We found in the analysis that the last hurdle to fall is typically hurdle (C), happening as frequently as all the other hurdles combined. An illustrative example is the history of developing a cure for hepatitis C. Hepatitis, or inflammation of the liver, has long been a part of human history. The symptoms can be severe, including abdominal pain, tiredness, jaundice (the yellowing of skin and eyes), and even liver failure and death.^96^ Patients with hepatitis C could be identified reliably from 1989^97,98^ and their responses measured quantitatively from 1993^99^ when diagnostics were developed that honed in on what was then known as non-A non-B hepatitis. Viral targets were known and accessible to small molecules from 1993,^100^ and yet, despite repeated efforts by companies such as Roche and Chiron, development proved fruitless. Unlike in many other infectious diseases, it was not possible to grow the pathogen in culture, leaving one key hurdle still in place. The wasted effort to cure hepatitis C before all the hurdles were overcome contributed to the poor performance and ultimate demise of Chiron Corporation as an independent entity—and of course left patients with a large, unmet need.^101,102^ The discovery of the replicon model in 1999^103^ proved to be the unlock, with—just a few years later, in 2003—Pharmasset running the screens that led to teleprevir,^104^ ushering in the era of direct-acting antivirals that specifically target hepatitis C. Today, cure rates for chronic hepatitis C are 90%—an incredible outcome for a previously chronic and sometimes fatal disease.^105^

The hepatitis C story is not unique and in fact is the most common pattern. According to our analysis, lack of good preclinical models remains the key stumbling block across diseases, with many models based on animals or biochemical assays having low correlation to the human organism.^106,107,108^ In particular, the models for chronic and progressive diseases, which represent the bulk of disease burden,^109^ often fail to recapitulate the slow, degenerative nature of these diseases.^107,108^ This has been particularly challenging in neuroscience, where animal models have even less correlation given the uniqueness of human brain expansion during development,^37^ and least challenging in infectious diseases where often only the pathogen needs to be modeled.^15^ This disparity has resulted in neuroscience typically having the lowest success rates in development, and infectious disease the highest.^15^

An emerging response from biopharmaceutical leaders when faced with this problem is to minimize the reliance on animal and assay-based models and focus more on what we can learn from human genetics:^23,24,25^ omics datasets properly coupled to longitudinal phenotypic data can reveal much about what can be important in disease,^110,111^ and when followed up with functional assessment of targets, can be a powerful way to identify and validate targets.^112,113^ This is indeed an appropriate response—one that has been responsible for recent improvements in productivity,^7,25^ and which, according to the findings in our current study, should very much be a focus of even deeper efforts going forward. It is a particularly appropriate approach for monogenic diseases, which should not be underestimated, as there are now more than 4,000 such identified conditions.^114^ However, it can be challenging for many other diseases, given the high polygenicity of many conditions and pleiotropy of the genes involved.^115^

Solving this issue—finding preclinical models that accurately predict human clinical response—is more important than is typically given credit by the industry.^39,116^ Even small changes in predictive value in preclinical can have large impacts on the total productivity, perhaps as much as increasing pipelines tenfold.^117,118^ So it behooves the biopharmaceutical industry and those that support it to make progress on this element of the drug development value chain that is so critical and yet currently so underdeveloped.

### The missing homunculus

In medieval literature, a homunculus was a miniature human created in a flask by alchemy.^119^ Imagine a perfect homunculus that fulfilled the following criteria: exact recapitulation of human clinical response, faster than real time, inexpensive, easy to use, and fully ethical to employ for drug testing. What would drug developers do differently? They would test much more rapidly and iteratively. Successes in the lab would guarantee successes in the clinic, and even failures would rapidly advance our understanding and contribute to better overall future success rates.

No preclinical system with the features of a perfect homunculus exists today, but its absence cannot yet be attributed to impossibility, given the relatively little effort that has been deployed to create one (see ahead). There are a range of possible avenues to explore to develop, if not a true homunculus, at least preclinical tests with much greater predictive value. These include organoid,^37,40,120,121,122^ organ-on-chip, body-on-chip,^123,124,125,126^ cell culture,^38,41,127,128^ explant,^129^ human phase zero,^116^ and in silico^130,131,132^ solutions. It is beyond the range of this paper to delve specifically into each of these or determine which might be most fruitful. Whatever is selected, there should be rigorous backtesting of both clinical failures and successes to determine the predictive value of the system(s).^133^

A corollary to creation and validation of models with predictive value is elimination of models with low predictive value. Too often, there is a dearth of backtesting such that it is unknown whether the models in current use are actually predictive. The predictions made by the models are not always even heeded, and yet they remain in use for no clear reason other than legacy, with positive results lauded, but negative results explained away.^134,135,136,137^

Despite the importance of better translational models, there is very limited activity to develop them. Academic labs have made initial forays, but have not typically developed models to full fruition. Pharmaceutical companies have much of their budgets deployed in asset-specific activities, and have not undertaken efforts of significant size to develop and back validate cross-program preclinical models. VC companies have made only extremely limited investments. Our own analysis finds that almost all VC investment is concentrated on hurdles (A) and (B), with less than 1% on hurdle (C) (see exhibit 5).

**Exhibit 5:**
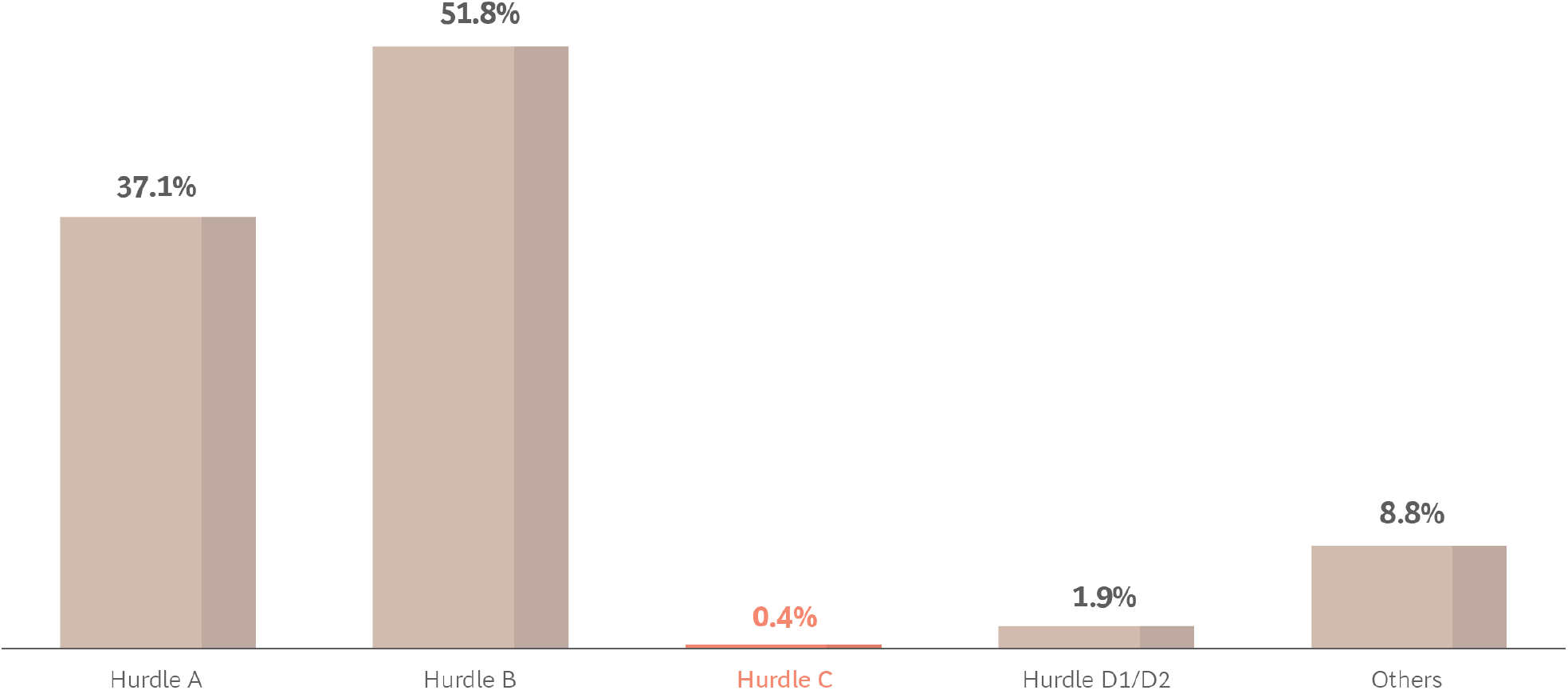
Percentage of VC-funded companies focused on each hurdle. In a sample of 838 VC-funded companies, we assessed the focus of activity, with the vast majority focused on hurdles (A) and (B), with very few on hurdle (C).

Our findings are corroborated by others who have found only 5% or less of funding going to companies focused on translational research,^138,139^ with very few startups active on the topic.^39,126^

The reason for this apparent market failure is that it historically has been difficult or impossible to make the kinds of financial returns from preclinical models (which are sold by the unit) that are available from the development of new modalities (which generally command a royalty stream from any product developed using the modality).^140,141^ This leaves a gap that is also an opportunity that could be addressed in multiple ways.

A private player could exploit this market imperfection by developing better translational models and using them in a proprietary way to pursue drug development with an advantage. A public-private partnership could be developed to deploy the best of industry, academia, and government to create one of these models,^142^ analogous to the work of the Accelerating Medicines Project (AMP) on target discovery.^143^ A consortium model could be used, building on nascent efforts at Transcelerate Biopharma’s BioCelerate subsidiary.^144^ Or government could take on the challenge directly, perhaps building on the National Center for Advancing Translational Sciences’s Tissue Chip Testing Centers and related efforts to build better translational models.^145^

## Concluding Remarks

We have seen the value of applying frameworks such as this one to individual program progression decisions in biopharma, resulting in an upswing in R&D productivity in recent years. It is our hope that applying this thinking at the level of diseases will help drive progress even more broadly by directing efforts in the most fruitful ways to overcome the challenges currently limiting progress in many diseases.

## Data Availability

Available data produced in the present work are contained in the manuscript.

## Author Contributions

MSR conceived the analysis; MSR and US oversaw the analysis; JD, MJD, MD, RAF, and SCH gathered data and conducted analyses; MSR and JD drafted the paper; MJD, MD, RAF, SCH, and US edited and approved the paper.

## Competing Interests

MSR, JD, and US are employees of Boston Consulting Group (BCG), a management consultancy that works with the world’s leading biopharmaceutical companies. MJD, MD, RAF, and SCH are previously employees of BCG. The research for this specific article was funded by BCG’s Health Care practice.

MSR is at Boston Consulting Group, Boston, Massachusetts, USA

JD is at Boston Consulting Group, Washington, D.C., USA

MJD is at Pfizer, New York, New York, USA

MD is at Eisai, Boston, Massachusetts, USA

RAF is at Handshake Health, Heidelberg, Germany

SCH is at BioNTech US, Cambridge, Massachusetts, USA

US is at Boston Consulting Group, Zurich, Switzerland

